# Examining the Prevalence and Risk Factors of Genital Warts among HIV-Infected and HIV-Negative Women: A Cross-Sectional Study in Cameroon

**DOI:** 10.1101/2024.04.03.24305297

**Authors:** Daina Charnelle Fougang, Martin Kuete

## Abstract

**Background:** Current research primarily focuses on high-risk strains of HPV associated with cervical cancer, overlooking risk factors for low-risk HPV infections like genital warts among vulnerable women, especially concerning their HIV serology. Understanding the interplay between genital warts, HIV status, and socio-cultural factors is crucial for informing targeted prevention to alleviate the burden of genital warts in vulnerable populations.

**Aims:** The purpose of this study is to determine the prevalence and risk factors of genital warts among women based on their HIV serostatus.

**Materials and Methods:** A cross-sectional study was conducted among women seeking gynecological consultation at a public Hospital in Cameroon. Data were collected through a survey and medical records, then analyzed using SPSS version 18.0.

**Results:** Among 257 women attending the hospital, 60 had genital warts, with the majority being HIV-positive (63.33%). Notably, 67% of these women sought gynecology consultation for the first time for genital warts. The location and types of genital warts were associated with HIV status, with papule genital warts being more prevalent among HIV-infected women and acuminate warts among non-infected women. Additionally, associations were found between smoking, multiple sexual partners, and genital warts among HIV-positive women. The reliance on traditional medicine or homemade remedies (85%) and the high prevalence of self-medication (75%) were also observed.

**Conclusions:** Addressing socio-cultural barriers is essential to enhance healthcare-seeking behaviour, facilitate early detection and treatment, and alleviate the burden of genital warts and HIV. Efforts should prioritize awareness-raising, enhancing healthcare access, and integrating traditional medicine into comprehensive healthcare systems.

## Background

Cervical cancer remains a significant public health challenge in Africa, with a high prevalence observed across the continent (Alhamlan et al., 2021).However, there is a notable gap in understanding the risk factors associated with low-risk HPV infections, such as genital warts, particularly among women with different HIV serostatus (Wei et al.).Genital warts are primarily caused by low-risk Human papillomavirus (HPV) types 6 or 11, accounting for approximately 90% of cases. These warts can have a significant impact on the well-being of patients, causing psychological and psychosexual distress, and leading to anxiety (O’Mahony et al., 2019). The regression of genital warts depends on the patient’s immunity. HPV infection is prevalent worldwide, particularly among people infected with the human immunodeficiency virus (HIV) (O’Mahony et al., 2019). HIV is a retrovirus that attacks the Cluster of Differentiating 4 (DC4) lymphocytes, leading to AIDS (acquired immunodeficiency syndrome) and making the body more vulnerable to other diseases or viruses such as HPV(Global, 2021) (Tartaglia et al., 2017). People with HIV are more vulnerable to potential HPV infection, which is also one of the most prevalent sexual virus infections in Cameroon [9]. The study indicated a high prevalence of HIV in Cameroon (43.1%) among women, with 8.02 million women at risk for cervical cancer [4,10]. However, developing countries face challenges with accessibility to both HPV vaccine awareness and antiretroviral (Tekbaş & Charnelle, 2023). The lack of knowledge about the social determinants of health, accessibility to appropriate care, and HPV vaccination in developing countries further exacerbates the burden of HPV-related infections, including genital warts, particularly among vulnerable populations such as those living with HIV(McDaniel et al., 2019; Tekbaş & Charnelle, 2023). In addition to medical factors, socio-cultural barriers play a crucial role in shaping the prevalence and management of HPV-related conditions (Peterson et al., 2022). Stigmatization surrounding sexually transmitted infections, including HPV, is pervasive, particularly among vulnerable populations such as HIV-infected individuals (Peterson et al., 2022). This stigma can act as a barrier to seeking appropriate medical care and may contribute to the underreporting of HPV-related symptoms. Additionally, despite the significant impact of genital warts on patients’ well-being, the research and attention in developing countries’ healthcare systems have been focused on cervical cancer, while genital warts have been relatively understudied and overlooked [6](Wei et al.). This gap underscores the need for more comprehensive studies and awareness campaigns that encompass the broader spectrum of HPV-related conditions, including genital warts(Peterson et al., 2022). Furthermore, traditional medicine practices often prevail in women’s healthcare in many African regions (Fantaye et al., 2019). The reliance on traditional remedies and treatments may reflect cultural beliefs, accessibility issues, or a lack of trust in conventional healthcare systems (M. R. Musie et al., 2022). Understanding the interplay between traditional and modern healthcare practices is essential for developing comprehensive approaches to HPV prevention and management. Given the significant importance of sociocultural factors in women’s health in Africa(Maurine R Musie et al., 2022), there is a need to identify exposure behaviour associated with genital warts specifically among women with varying HIV serology. Therefore, the purpose of this study is to determine the prevalence and risk factors of genital warts among women based on their HIV serostatus. By elucidating the relationship between genital warts, HIV status, and socio-cultural factors, to inform targeted prevention and management strategies to mitigate the burden of genital warts in vulnerable populations.

## 1. Materials and methods

### 2.1. The study designs

This study employed a cross-sectional design and was conducted at the gynaecological consultation service at a public hospital in the central region of Cameroon.

### 2.2. Data collection

The study included women aged 18 years and above diagnosed with genital warts. Consecutive non-probability sampling was utilized, with 60 out of the 257 women who received gynaecological consultations between January and October 2023 being included in the study due to their genital wart diagnosis.

#### 2.2.1. Questionnaire form

A questionnaire was developed by the researchers based on recommendations and literature on HPV infections in developing countries. It focused on sociodemographic information, HIV status, location and type of genital warts, and associated behaviours and factors.

#### 2.2.3. Data Collection Procedure

Information on HIV serology status and clinical characteristics of genital warts was obtained through a combination of medical record review and clinical examination conducted by a gynecologist. key areas, including HIV status, the location and type of genital warts, and factors associated with condyloma. Additionally, socio-demographic data, behavoir, and factors associated with the condition were gathered through patients using the developed questionnaire.

### 2.3. Evaluation of research data

The collected data were analyzed using SPSS version 18.0 software. Descriptive statistics such as percentages, frequencies, means, and standard deviations were utilized to analyze the data. Parametric tests, including one-way ANOVA, and non-parametric tests, such as Kruskal-Wallis test, were employed to analyze relationships between variables.

### 2.4. Ethical considerations

The research received proper authorization and ethics clearance from the Ethics Committee of Universite des Montagnes de Bagante (authorization IBR No 152/UDM/PR/CIE) and the Institutional Ethics Committee of Human Health of the hospital. Participants were provided with information regarding anonymity, confidentiality, and their rights, and free and informed consent was obtained from each participant.

## 3. Results

### 3.1. Profiles of participants

The results indicate that among the 60 participants diagnosed with genital warts, the mean age was 26 years old. The age range varied from 15 to 62 years old, with the youngest participant being 15 years old and the oldest being 62 years old. Nearly half of the women fell into the age bracket of 26-30 years old (49.8%), and a majority of them were single (58.33%). Furthermore, the majority of participants had attained a university level of education (41.1%), and 88.33% reported having at least three children (see *Table 1*).

**Table 1.** Socio-demographic characteristics of women.

| Descriptive characteristics |  | n | % |
| --- | --- | --- | --- |
| Number of children | 0-3 | 53 | **88.33** |
|  | 4-7 | 3 | 5.00 |
|  | More than 7 | 4 | 6.66 |
|  | 15-25 | 30 | 53.00 |
| Age | 26-30 | 106 | **49.80** |
|  | Over 31 | 22 | 10.30 |
|  | Primary | 16 | 26.70 |
| Education level | High school | 19 | 31.70 |
|  | University | 25 | **41.70** |
| **Marital status** | Single | 35 | **58.33** |
|  | Couple/boy-girlfriend | 4 | 6.66 |
|  | Married | 20 | 33.30 |
|  | Polygamic | 1 | 1.66 |

### 3.2. The Prevalence of genital warts among women with HIV-positive and negative

In this study, out of 257 women attending gynaecologist consultations, 60 of them (23%) were diagnosed with genital warts. Among the women with genital warts, 38 (63.33%) were HIV-positive, while 22 (36.6%) were HIV-negative (p=0.039).

**Table 2** sheds light on the care and management of genital warts, focusing on the seeking behaviour among participants. The majority noticed their genital warts themselves (60%), with most of them having their first gynaecologist consultation after noticing warts (67.14%). A significant proportion had not visited a gynaecologist in over 3 months (80%). Furthermore, a considerable number used traditional medicine or homemade remedies (85%) and engaged in self-medication (75%).

**Table 2:** Genital Warts Care and Management: seeking behavior among Women in Cameroon.

| **Questions** | **Responses n (%)** |
| --- | --- |
| Who noticed your genital warts first? |  |
| Self | 36 (60.%) |
| Partner | 10 (16.67%) |
| Doctor | 14 (23.33%) |
| Is it your first time coming for a gynaecologist consultation since having genital warts? |  |
| Yes | 47 (67.14%) |
| No | 23 (32.86%) |
| Was your last gynaecologist consultation date more than 3 months ago? |  |
| Yes | 48 (80%) |
| No | 12 (20%) |
| Do you use traditional medicine or homemade remedies for your genital warts? |  |
| Yes | 51(85%) |
| No | 09 (15.00%) |
| Did you engage in self-medication upon noticing genital warts? |  |
| Yes | 45(75%) |
| No | 15 (25.%) |

### 3.3. Characteristic of genital warts among women with HIV-positive and negative

In terms of the localization of genital warts, a statistically significant association (p≤0.05) was observed between the vulva and vaginal sites and HIV serology (*Table 3*). The genital warts were predominantly localized on the vagina (60%), with the vulva being the second most common site (35%). Notably, vaginal presentation was more frequently observed among HIV-infected women (63.2%) compared to HIV-negative women (54.5%) (*Table 3*). Women who were HIV-negative were more likely (0.90±0.31) to have condyloma on the vulvar compared to HIV-positive women (0.85±0.36).

**Table 3.** Repartition of the characteristics of genital warts in the two groups.

| Characteristics | HIV (+) |  | HIV (-) |  | *p* |
| --- | --- | --- | --- | --- | --- |
| | n (%) | (mean ±SD) | n (%) | (mean ±SD) | |
| **localization** |  |  |  |  |  |
| Vulva | 12(31.6) | **0.85±0.36** | 9(40.9) | **0.90±0.31** | **0.037\*** |
| Lips | 1(2.6) | 0.70±0.26 | | 1.15±0.5 | 0.514 |
| Anal | 1(2.6) | 0.7±0.26 | 1(4.5) | 0.1±0.36 | 0.665 |
| Vagina | **24 (63.2)** | **0.63±0.48** | 0(0.00) | **0.54±0.5** | **0.017\*** |
| **Type of genital warts** |  |  | 12(54.5) |  |  |
| | | 0.52±0.5 | | 0.59±0.5 | 0.118 |
| Acuminate | 20(52.6) | **0.9±0.29** |  | **0.21±0.41** | **0.014\*** |
| Papule | 8(21.1) | 0.26±0.44 | 13(59.1) | 0.31±0.47 | 0.187 |
| Flat | 10(26.30) |  | 2(9.10) |  |  |
|  |  |  | 7(31.8) |  |  |

ANOVA test, \* $p < 0.05$

Regarding the type of genital warts, there was a statistically significant association (p≤0.05) between papule genital warts and HIV serology. Indeed, HIV-positive women were more inclined (0.9±0.29) to present papule genital warts compared to non-infected women (0.21±0.41). Additionally, acuminate genital warts were found to be more prevalent overall (55%). However, non-infected women exhibited a lower incidence of acuminate genital warts (52.6%) compared to the HIV-negative group (59.1%) *(Table 3*).

### 3.5. The associated factors of genital warts among the two groups

The most prevalent risk factors in the HIV-positive group were smoking (70.8%) and having multiple sexual partners (60.5%). Conversely, among women with HIV-negative status, the most common risk factors were smoking (66.7%) and early intercourse (59.1%). A statistically significant association (p=0.00) was found between smoking and HIV-positive serology as a risk factor. However, smoking was not significantly associated with negative HIV serology in women (p>0.05). Furthermore, there was a statistically significant association between having multiple sexual partners and being HIV-positive (Table 4).

**Table 4.**
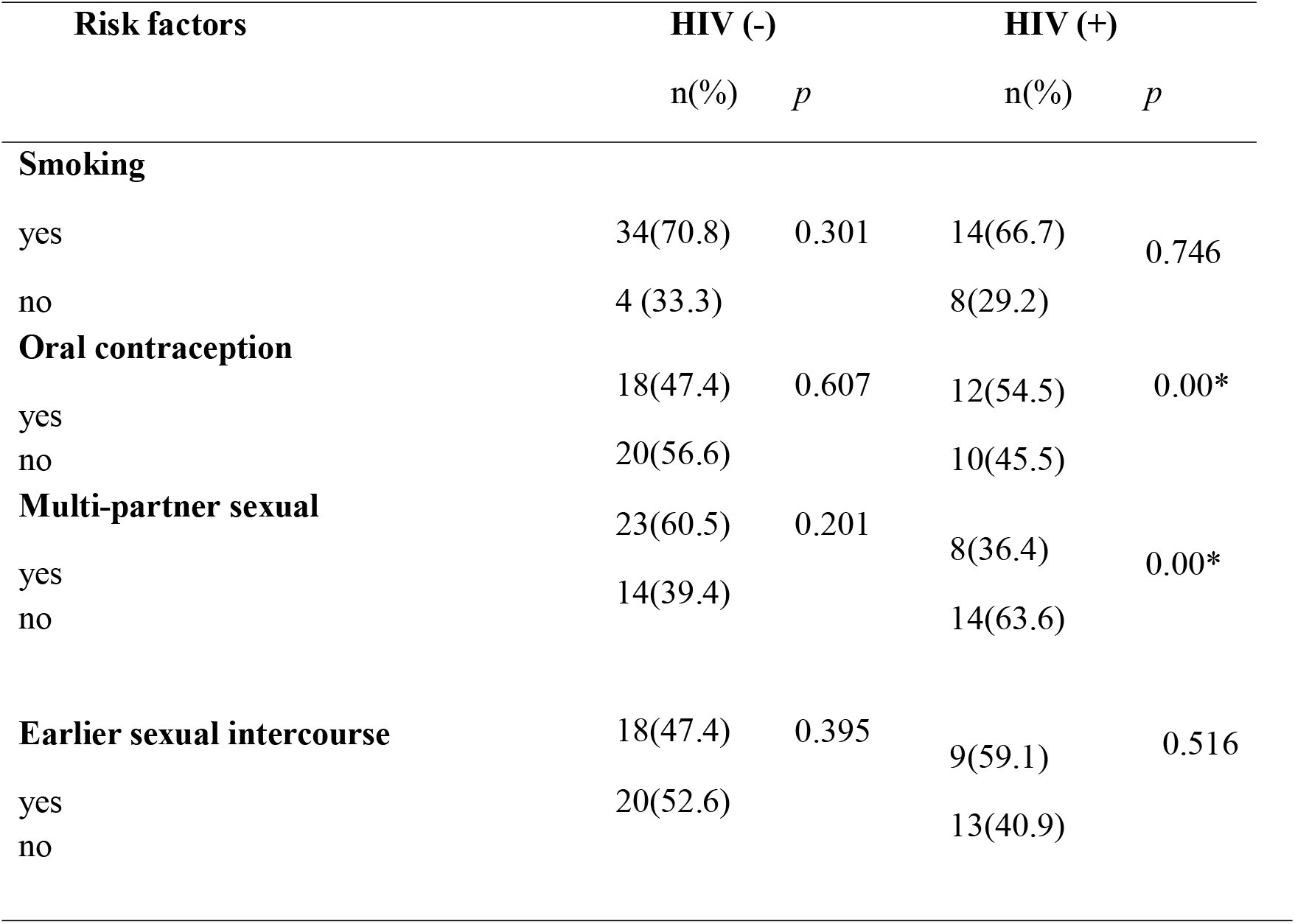
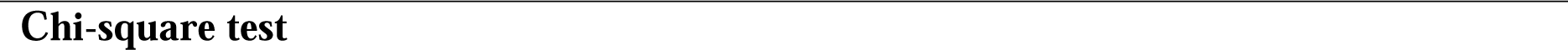
Repartition of the associated factor of genital warts in the two groups.

## 4. Discussion

The high prevalence of genital warts among participants in this study, especially among women aged 26 to 30, underscores the urgency for targeted prevention and screening efforts in this age group. This prevalence aligns with findings from similar studies in developing countries(Chikandiwa et al., 2018; Simo et al., 2021), emphasizing the need for tailored interventions. Indeed, this could be due to sexual activity being more frequent among women in this age group, leading to a higher risk of exposure to HPV(Simo et al., 2021). The higher prevalence among educated women may reflect increased health-seeking behaviour and awareness of preventive measures like HPV vaccination and cervical cancer screening (Chikandiwa et al., 2018; Simo et al., 2021; Wabo et al., 2019). The association between higher parity and increased risk of HPV infection corroborates previous research (Tekalegn et al., 2022). Factors such as changes in the cervix during pregnancy and childbirth may contribute to this association. Additionally, being single may indicate a higher likelihood of engaging in unprotected sexual activity, highlighting broader social and economic factors at play, such as barriers to healthcare access and socioeconomic status. Targeted prevention and screening efforts are crucial to address the high prevalence of genital warts among Cameroonian women. Moreover, addressing broader social and economic factors is essential to ensure equitable access to healthcare and mitigate the burden of HPV-related conditions.

### 4.1. The Genital wart among women HIV positive and HIV negative

This study demonstrates a significant association between HIV serology and genital warts among women in Cameroon, indicating that HIV-positive individuals are at greater risk of developing genital warts. HIV-induced immune regression is a major risk factor for HPV infection, exacerbating HPV-related symptoms (Sosso et al., 2020; Zayats et al., 2022). The heightened prevalence of genital warts among HIV-positive women underscores the elevated cervical cancer rates observed in Africa (Ba et al., 2021). Notably, HIV-positive individuals exhibit a higher prevalence of high-risk HPV, highlighting HIV as a potential risk factor for genital cancer (Dreyer et al., 2022; Hlahla, 2023; Riddell IV et al., 2022; Sosso et al., 2020). The weakened immune system in HIV facilitates HPV replication and progression, leading to cervical intraepithelial lesions and genital warts (Taku et al., 2020). Study indicated HIV target cells are found to be highly concentrated in the outermost skin layer of anogenital warts, providing a possible explanation for the observed association between HIV and genital warts (S. B. Dhumale et al., 2017). Additionally, a recent study demonstrated that although low-risk HPV co-expression is not directly associated with genital cancer, it can increase DNA damage due to the accumulation of somatic mutations(Pudney et al., 2019; Uehara et al., 2021). The observed association between HPV and HIV infections necessitates comprehensive prevention and management strategies for both conditions, particularly among vulnerable populations in developing countries.

### 4.2. Characteristic of genital warts among women with negative HIV and positive

This study reveals distinct characteristics of genital warts among women with positive and negative HIV status. Vaginal localization of genital warts is more prevalent among HIV-positive women, whereas vulvar localization is more common among HIV-negative women. This disparity may be attributed to differences in sexual behavior following counseling and screening, with HIV-positive individuals adopting safer sexual practices. This finding is consistent with a previous study that also reported higher rates of genital warts in the vaginal area among HIV-positive women (S. B. Dhumale et al., 2017). Interestingly, a link was observed between positive HIV serology and vaginal condyloma, while negative HIV serology was associated with vulvar localization (p = 0.037). This disparity may be attributed to differences in sexual behaviour following counselling and screening, with HIV-positive individuals adopting safer sexual practices. In this study, acuminate genital warts were more frequently found in HIV-negative women and may regress with a normal immune response, whereas papule genital warts, which were frequent among HIV-infected women could be less likely to regress (Pudney et al., 2019). Although there are limited studies comparing the types of genital warts in both groups, the results suggest that papule genital warts affect the mucosa more, which is in close proximity to low-grade cervical intraepithelial neoplasia (Shashikant Balakrishana Dhumale et al., 2017; Kosz et al., 2020).

### 4.2. The risk associated with HPV infection

Social determinants of health significantly shape the risk associated with HPV and HIV infections in developing countries. This study identifies distinct risk factors among HIV-positive and HIV-negative women, with sexual behaviour emerging as a primary risk factor. Among HIV-positive women, early oral contraceptive use emerged as a prominent risk factor, possibly due to its impact on hormonal concentrations, leading to an increased risk of genital warts (Kosz et al., 2020). Early oral contraceptive use is prominent among HIV-positive women, while smoking and multiple sexual partners are significant risk factors among HIV-negative women (Sarma et al., 2023). The observed association in this study between smoking, multiple sexual partners, and genital warts among HIV-positive women underscores the complex interplay between HPV and HIV infections [21]. The high prevalence of genital warts and HIV among these women underscores the urgency of understanding and addressing these issues within the context of socio-cultural factors. Firstly, the majority of women noticed their genital warts themselves (60%), indicating a proactive approach to their health. However, this also suggests a potential lack of awareness about preventive measures or the importance of regular gynecologist consultations. Moreover, most women sought gynaecologist consultation only after noticing warts (67.14%), indicating a reactive rather than proactive approach to healthcare. This delay in seeking medical attention could be attributed to various factors, including cultural taboos, fear of stigma, or limited access to healthcare facilities. Additionally, a significant proportion of women had not visited a gynaecologist in over 3 months (80%). This highlights a lack of consistent gynaecological care, which may be influenced by socio-economic factors such as financial constraints, transportation issues, or cultural beliefs regarding healthcare utilization. The reliance on traditional medicine or homemade remedies by a considerable number of women (85%) further emphasizes the influence of socio-cultural beliefs on healthcare-seeking behaviour. Traditional medicine may be perceived as more accessible, affordable, or culturally acceptable, leading to its widespread use despite potential risks (Organization, 2015). Moreover, the high prevalence of self-medication (75%) among women reflects the limited access to healthcare services or the lack of trust in the healthcare system. Self-medication can lead to improper treatment, delayed diagnosis, or adverse health outcomes, highlighting the need for improved access to quality healthcare services and public health education initiatives.

Addressing socio-cultural barriers is crucial in improving healthcare-seeking behavior, promoting early detection and treatment, and reducing the burden of genital warts and HIV. Efforts should focus on raising awareness, improving access to healthcare services, and integrating traditional medicine practices into comprehensive healthcare systems. Addressing social determinants of health in prevention strategies can effectively lower HPV and HIV incidence in developing countries, ultimately improving the health outcomes of vulnerable populations (Osazuwa-Peters et al., 2019).

## Limitations and recommendation

This study was conducted in a single region of Cameroon and may not be generalizable to other regions or countries. Future studies could assess the psychological and social impact of genital warts on women in Cameroon and explore ways to improve.

## Data Availability

All data produced in the present study are available upon reasonable request to the authors

## Acknowledgements

We are thankful to all the women who agreed to participate and answer our interviews.

## Availability of data and materials

The authors declare that all data supporting the findings of this study are available on request due to privacy/ethical restrictions.

## References

Alhamlan, F. S., Alfageeh, M. B., Al Mushait, M. A., Al-Badawi, I. A., & Al-Ahdal, M. N. (2021). Human papillomavirus-associated cancers. Microbial Pathogenesis: Infection and Immunity, 1–14.

Ba, D. M., Ssentongo, P., Musa, J., Agbese, E., Diakite, B., Traore, C. B., Wang, S., & Maiga, M. (2021). Prevalence and determinants of cervical cancer screening in five sub-Saharan African countries: a population-based study. Cancer epidemiology, 72, 101930.

Chikandiwa, A., Kelly, H., Sawadogo, B., Ngou, J., Pisa, P. T., Gibson, L., Didelot, M. N., Meda, N., Weiss, H. A., Segondy, M., Mayaud, P., & Delany-Moretlwe, S. (2018). Prevalence, incidence and correlates of low risk HPV infection and anogenital warts in a cohort of women living with HIV in Burkina Faso and South Africa. PLoS One, 13(5), e0196018. 10.1371/journal.pone.0196018

Dhumale, S. B., Sharma, S., & Gulbake, A. (2017). Ano-Genital Warts and HIV Status-A Clinical Study. J Clin Diagn Res, 11(1), Wc01–wc04. 10.7860/jcdr/2017/24610.9171

Dhumale, S. B., Sharma, S., & Gulbake, A. (2017). Ano-genital warts and HIV status–A clinical study. Journal of Clinical and Diagnostic Research: JCDR, 11(1), WC01.

Dreyer, G., Snyman, L. C., Van der Merwe, F., Richter, K. L., Dreyer, G., Visser, C., & Botha, M. (2022). Phase I of the DiaVACCS screening trial: Study design, methods, population demographics and baseline results. South African Medical Journal, 112(7), 478–486.

Fantaye, A. W., Gunawardena, N., & Yaya, S. (2019). Preferences for formal and traditional sources of childbirth and postnatal care among women in rural Africa: A systematic review. PLoS One, 14(9), e0222110. 10.1371/journal.pone.0222110

Global, H. (2021). AIDS statistics—2019 fact sheet [https://www.unaids.org/en/resources/fact-sheet]. In: Accessed.

Hlahla, K. (2023). Risk factors associated with the presence of cervical lesions in women attending a family planning clinic in Harare Zimbabwe: a cross-sectional study.

Kosz, K., Zielińska, M., Kuchnicka, A., Zarankiewicz, N., & Ciseł, B. (2020). Relationship between HPV and HIV. Prevalence, molecular mechanisms and screening of HPV among HIV infected women. Journal of Education, Health and Sport, 10(7), 127–137. 10.12775/JEHS.2020.10.07.013

McDaniel, J. T., Nuhu, K., Ruiz, J., & Alorbi, G. (2019). Social determinants of cancer incidence and mortality around the world: an ecological study. Global health promotion, 26(1), 41–49.

Musie, M. R., Anokwuru, R. A., Ngunyulu, R. N., & Lukhele, S. (2022). African indigenous beliefs and practices during pregnancy, birth and after birth. Working with indigenous knowledge: Strategies for health professionals [Internet].

Musie, M. R., Peu, M. D., & Bhana-Pema, V. (2022). Culturally appropriate care to support maternal positions during the second stage of labour: Midwives’ perspectives in South Africa. Afr J Prim Health Care Fam Med, 14(1), e1–e9. 10.4102/phcfm.v14i1.3292

O’Mahony, C., Gomberg, M., Skerlev, M., Alraddadi, A., de Las Heras-Alonso, M. E., Majewski, S., Nicolaidou, E., Serdaroğlu, S., Kutlubay, Z., Tawara, M., Stary, A., Al Hammadi, A., & Cusini, M. (2019). Position statement for the diagnosis and management of anogenital warts. J Eur Acad Dermatol Venereol, 33(6), 1006–1019. 10.1111/jdv.15570

Organization, W. H. (2015). WHO recommendations on partnership with traditional birth attendants. The WHO Reproductive Health Library, Geneva.

Osazuwa-Peters, N., Adjei Boakye, E., Rohde, R. L., Ganesh, R. N., Moiyadi, A. S., Hussaini, A. S., & Varvares, M. A. (2019). Understanding of risk factors for the human papillomavirus (HPV) infection based on gender and race. Scientific reports, 9(1), 1–7.

Peterson, C. E., Dykens, J. A., Weine, S. M., Holt, H. K., Fleurimont, J., Hutten, C. G., Wieser, J., Abuisneineh, F., Awadalla, S., Ongtengco, N. P., Gastala, N., & Jasenof, I. G. (2022). Assessing the interrelationship between stigma, social influence, and cervical cancer prevention in an urban underserved setting: An exploratory study. PLoS One, 17(12), e0278538. 10.1371/journal.pone.0278538

Pudney, J., Wangu, Z., Panther, L., Fugelso, D., Marathe, J. G., Sagar, M., Politch, J. A., & Anderson, D. J. (2019). Condylomata Acuminata (Anogenital Warts) Contain Accumulations of HIV-1 Target Cells That May Provide Portals for HIV Transmission. J Infect Dis, 219(2), 275–283. 10.1093/infdis/jiy505

Riddell IV, J., Brouwer, A. F., Walline, H. M., Campredon, L. P., Meza, R., Eisenberg, M. C., Andrus, E. C., Delinger, R. L., Yost, M. L., & McCloskey, J. K. (2022). Oral human papillomavirus prevalence, persistence, and risk-factors in HIV-positive and HIV-negative adults. Tumour Virus Research, 13, 200237.

Sarma, P., Barmon, D., Rai, A. K., Kataki, A. C., Sarma, A., Kakoti, L., Barman, D., & Kalita, M. (2023). Tobacco chewing, Alcohol consumption, reuse of cloth sanitary pads is significant risk factors for High Risk HPV infection and PAP positivity among Rural Women of Kamrup District, North-East India. medRxiv, 2023.2001. 2016.23284644.

Simo, R. T., Kiafon, F., Nangue, C., Goura, A. P., Ebune, J. L., Usani, M. C., Kamdje, A. H. N., Etet, P. F. S., & Telefo, P. B. (2021). Influence of HIV infection on the distribution of high-risk HPV types among women with cervical precancerous lesions in Yaounde, Cameroon. International Journal of Infectious Diseases, 110, 426–432.

Sosso, S. M., Tchouaket, M. C. T., Fokam, J., Simo, R. K., Torimiro, J., Tiga, A., Lobe, E. E., Ambada, G., Nange, A., & Semengue, E. N. J. (2020). Human immunodeficiency virus is a driven factor of human papilloma virus among women: evidence from a cross-sectional analysis in Yaoundé, Cameroon. Virology journal, 17, 1–7.

Taku, O., Businge, C. B., Mdaka, M. L., Phohlo, K., Basera, W., Garcia-Jardon, M., Meiring, T. L., Gyllensten, U., Williamson, A.-L., & Mbulawa, Z. Z. (2020). Human papillomavirus prevalence and risk factors among HIV-negative and HIV-positive women residing in rural Eastern Cape, South Africa. International Journal of Infectious Diseases, 95, 176–182.

Tartaglia, E., Falasca, K., Vecchiet, J., Sabusco, G. P., Picciano, G., Di Marco, R., & Ucciferri, C. (2017). Prevalence of HPV infection among HIV-positive and HIV-negative women in Central/Eastern Italy: Strategies of prevention. Oncol Lett, 14(6), 7629–7635. 10.3892/ol.2017.7140

Tekalegn, Y., Sahiledengle, B., Woldeyohannes, D., Atlaw, D., Degno, S., Desta, F., Bekele, K., Aseffa, T., Gezahegn, H., & Kene, C. (2022). High parity is associated with increased risk of cervical cancer: Systematic review and meta-analysis of case–control studies. Women’s Health, 18, 17455065221075904.

Tekbaş, S., & Charnelle, D. (2023). Knowledge, Behaviours and Affecting Factors About Human Papilloma Virus and Vaccination Among University Students [Üniversite Öğrencilerinin Human Papillomavirüs ve Aşisi Hakkindaki Bilgi, Davranişlari ve Etkileyen Faktörler]. Ordu Üniversitesi Hemşirelik Çalişmalari Dergisi, 6(3), 731–738. 10.38108/ouhcd.1170908

Uehara, K., Tanabe, Y., Hirota, S., Higa, S., Toyoda, Z., Kurima, K., Kina, S., Nakasone, T., Arasaki, A., & Kinjo, T. (2021). Co-expression of low-risk HPV E6/E7 and EBV LMP-1 leads to precancerous lesions by DNA damage. BMC cancer, 21(1), 1–13.

Wabo, B., Nsagha, D. S., Nana, T., Pokam, B. D. T., Njiomenie, G. F., Guemdjom, W. P., & Assob, J. C. N. (2019). Knowledge on cervical cancer and screening tests among women at two reference hospitals in Yaounde, Cameroon. International Journal of Biological and Chemical Sciences, 13(3), 1487–1495.

Wei, F., Georges, D., Man, I., Baussano, I., & Clifford, G. Causal Attribution of Human Papillomavirus Types to Invasive Cervical Cancer Worldwide: A Systematic Analysis of the Global Literature. Available at SSRN 4692586.

Zayats, R., Murooka, T. T., & McKinnon, L. R. (2022). HPV and the Risk of HIV Acquisition in Women. Frontiers in Cellular and Infection Microbiology, 6.

